# Extract-Free One-Pot Ambient RPA-CRISPR Detection of *Plasmodium* in Whole Blood

**DOI:** 10.64898/2026.03.17.26348654

**Authors:** Jipei Liao, Yun Su, Feng Jiang

## Abstract

**Background:** Malaria caused by *Plasmodium* is a major global health burden. Rapid and accurate malaria detection is essential for effective disease control. However, most molecular diagnostics require DNA extraction, thermal cycling, and specialized infrastructure, limiting use in resource-limited settings. This study aimed to develop an extraction-free, one-pot CRISPR-based platform for rapid *Plasmodium* detection directly from whole blood under ambient conditions.

**Methods:** Crude blood lysates were directly applied to a one-pot recombinase polymerase amplification (RPA)-CRISPR reaction coupled with lateral flow readout. Analytical sensitivity and specificity were evaluated using purified DNA and whole-blood lysates. Diagnostic performance was assessed using a clinical cohort comprising 116 malaria cases and 109 negative controls.

**Results:** The extract-free RPA-CRISPR assay achieved an analytical limit of detection of 100 copies/µL from crude blood lysates, corresponding approximately to 12-20 parasites/µL. No cross-reactivity was observed across *Plasmodium* species. In clinical evaluation, the assay demonstrated strong concordance with reference PCR, achieving 93.1% sensitivity and 100% specificity. Sensitivity varied with parasite burden, reaching 97.8% for high-density infections, 95.1% for moderate density, and 82.8% for low-density infections. The whole workflow was completed within 40 minutes at room temperature without specialized instrumentation.

**Conclusions:** An extract-free, one-pot ambient-temperature RPA-CRISPR platform allows rapid detection of *Plasmodium* directly from whole blood with high specificity and sensitivity across parasite burdens, supporting its potential for decentralized malaria diagnostics. Larger multicenter field studies are needed to validate performance under real-world conditions.

## INTRODUCTION

Malaria, caused by protozoan parasites of the genus *Plasmodium*, remains a major global public health challenge ^1^. Approximately 263 million malaria cases and 597,000 deaths occur worldwide each year ^1^. Five *Plasmodium* species are known to cause malaria in humans: *Plasmodium falciparum (P. falciparum), Plasmodium vivax (P. vivax), Plasmodium ovale, Plasmodium malariae*, and *Plasmodium knowlesi*. Among these, *P. falciparum* is responsible for most severe malaria cases and malaria-related mortality globally, whereas *P. vivax* is the most geographically widespread species and a major contributor to recurrent malaria due to its ability to form dormant liver hypnozoites^2^. Because malaria frequently presents nonspecific clinical manifestations such as fever, chills, and influenza-like symptoms, timely and accurate diagnosis of malaria is essential to reduce morbidity and mortality and to limit the emergence of antimalarial drug resistance^1^. Current diagnostic approaches include microscopic examination of Giemsa-stained blood smears, rapid diagnostic tests (RDTs), and nucleic acid-based detection methods such as polymerase chain reaction (PCR) or molecular panels, including the BioFire Global Fever Panel ^3^. While microscopy remains the reference standard, it requires trained personnel and may lack sensitivity at low parasitemia^4^. RDTs offer rapid results but can exhibit reduced sensitivity, particularly in low-density infections or in cases involving non-falciparum species^5^. PCR-based assays provide high analytical sensitivity but require DNA extraction, thermal cycling, and specialized laboratory infrastructure, limiting their deployment in decentralized or resource-limited settings^6^. Therefore, there is a critical need for diagnostic tools that combine molecular-level sensitivity with operational simplicity and field compatibility.

Clustered Regularly Interspaced Short Palindromic Repeats (CRISPR) are adaptive immune systems found in bacteria and archaea that protect against invading genetic elements^7^. CRISPR-associated (Cas) proteins, guided by CRISPR RNA (crRNA), enable sequence-specific recognition and cleavage of nucleic acids ^8^. This property has made CRISPR-Cas a powerful platform for sensitive and sequence-specific nucleic acid detection^8^. We have shown that CRISPR-Cas12a detection platforms achieve PCR-comparable analytical sensitivity while significantly reducing workflow complexity for the detection of viral and DNA targets ^9-14^. CRISPR-based systems have recently demonstrated high sensitivity for detecting *Plasmodium* species in clinical specimens ^15,16^. Yet, the CRISPR workflows require multi-step DNA extraction and incubation at controlled temperatures (typically 42 °C), creating dependence on electricity and heating equipment that limits true point-of-care (POC) deployment^17^. Recent studies have demonstrated room-temperature RPA-CRISPR detection using optimized Cas12a orthologs^18-21^. However, most reported implementations still rely on purified nucleic acids, multi-step processing, or laboratory-prepared sample inputs, limiting their practical deployment in true POC settings^18-21^. Furthermore, few studies have addressed direct detection from crude whole blood matrices without heating or centrifugation^22,23^. For malaria diagnostics, where field implementation is essential, eliminating both thermal control and nucleic acid extraction remains a critical unmet need. To address this clinical need, we aimed to develop a one-pot, CRISPR diagnostic platform for the rapid, extraction-free detection of *Plasmodium* species directly from whole blood at room temperature. Our developed platform for *Plasmodium* detection maintains high analytical sensitivity and diagnostic accuracy, supporting its potential as a POC tool for decentralized and resource-limited settings.

## MATERIALS AND METHODS

### Overall Research Design

The workflow integrates three sequential components: (1) Extract-free chemical lysis of raw blood without heating or centrifugation; (2) a closed-tube, one-pot reaction combining recombinase polymerase amplification (RPA) and Cas12a-mediated CRISPR detection at 25 °C; and (3) visual readout via lateral flow assay (LFA). Analytical performance was first established using quantified control DNA templates for *Plasmodium* species to determine specificity and limit of detection. Clinical performance was subsequently assessed using patient whole-blood samples to evaluate diagnostic sensitivity, specificity, and overall concordance.

### Control *Plasmodium* DNA Templates

This study focused on the two most clinically significant human malaria species, *P. falciparum* and *P. vivax*. Genomic DNA from *P. falciparum* strain 3D7 (ATCC PRA-405D) and synthetic *P. vivax* DNA standards (ATCC PRA-3004SD) were obtained from the American Type Culture Collection (ATCC, Manassas, VA). The *P. falciparum* genomic DNA preparation represents the well-characterized 3D7 laboratory strain, whereas the

*P. vivax* synthetic DNA standard contains defined target regions of the conserved 18S rRNA gene. DNA concentrations were verified upon receipt and quantified using PCR to ensure accurate copy number determination. Stock solutions were aliquoted and stored at −20 °C to prevent freeze-thaw degradation. Serial dilutions were prepared in nuclease-free water for intrinsic analytical sensitivity testing or spiked into malaria-negative whole blood for matrix-based extract-free evaluations. Both genomic DNA preparations were used for the RPA-CRISPR assay optimization, analytical sensitivity testing, and species-specific validation experiments.

### Ambient-Temperature Extract-Free Sample Preparation

We adapted extract-free chemical lysis strategies ^24-26^ using a mild non-ionic detergent (Triton X-100; Sigma-Aldrich, St. Louis, MO, USA) at room temperature with minor modification. Five µL of EDTA-anticoagulated whole blood was mixed with 10 µL of lysis buffer containing 1.0% Triton X-100 in 1× phosphate-buffered saline (PBS; Thermo Fisher Scientific, Waltham, MA, USA; pH 7.4) supplemented with 0.1% bovine serum albumin (BSA; Sigma-Aldrich, St. Louis, MO, USA), followed by incubation at 25 °C for 10 minutes. No heating, centrifugation, or purification steps were performed. An aliquot (2-5 µL) of the resulting crude lysate was directly introduced into the one-pot RPA-Cas12a reaction.

### One-Pot RPA-CRISPR Reaction

A single-tube reaction integrating RPA with CRISPR-mediated detection was conducted using LbCas12a (Integrated DNA Technologies, Inc. IDT, Coralville, IA) as previously described ^20^. Lyophilized TwistAmp® Basic RPA pellets (TwistDx Ltd., Maidenhead, United Kingdom) were rehydrated according to the manufacturer’s instructions and combined in a single 15 µL reaction mixture containing forward and reverse primers targeting the conserved 18S rRNA gene of *P. falciparum* and *P. vivax*, pre-assembled Cas12a-crRNA ribonucleoprotein complexes specific to each species, a FAM-biotin-labeled single-stranded DNA reporter for lateral flow detection, dNTPs, and reaction buffer components (Sigma-Aldrich). Magnesium acetate (Sigma-Aldrich) was added immediately prior to incubation to initiate amplification. Amplification and CRISPR detection proceeded simultaneously within the same closed tube at 25 °C for 40-60 minutes. Corresponding primer for RPA and crRNA sequences are listed in Supplementary Table S1.

### Lateral Flow Assay (LFA) Readout

Following ambient incubation of the one-pot RPA-CRISPR reaction, visual detection was performed using commercially available lateral flow strips (HybriDetect Universal LFA Kit. Milenia Biotec GmbH, Giessen, Germany) as described in our previous reports ^9-14^. The reaction mixture contained a FAM-biotin-labeled single-stranded DNA reporter designed such that, upon target recognition and activation of Cas12a, collateral trans-cleavage of the reporter separated the FAM and biotin moieties, enabling capture of cleaved fragments at the test line. After 20 minutes of incubation at 25 °C, the lateral flow strip was inserted directly into the reaction tube. Cleavage of the reporter produced a visible test band for 1-5 minutes, while the control line confirmed proper flow and reagent integrity. The complete workflow—from crude blood lysis to visual interpretation—was accomplished in under 40 minutes at constant room temperature. Images of the developed strips were captured using a standard mobile phone camera for documentation and record keeping ^9-14^.

### Two-step RPA-CRISPR Assay

For the conventional two-step RPA-CRISPR assay, amplification and CRISPR detection were performed sequentially rather than in a single closed tube as described in our previous studies ^9-14^. Briefly, RPA amplification was first conducted using the same primer sets and reaction conditions described for the one-pot assay at 40 °C. After amplification (20-30 minutes), an aliquot (2-5 µL) of the RPA product was transferred to a separate tube containing the pre-assembled Cas12a-crRNA complex and reporter for CRISPR-mediated detection. The detection reaction proceeded at 40 °C for an additional 20-30 minutes, followed by lateral flow readout as described above.

### Preclinical Evaluation Using Blood Samples

EDTA-anticoagulated whole blood samples were obtained at Jiangsu Province Hospital of Chinese Medicine. All procedures were conducted under institutional review board-approved protocol, following the ethical standards. The clinical cohort comprised 225 specimens, including 116 PCR-confirmed malaria cases and 109 PCR-negative controls. Among the malaria cases, 65 were *P. falciparum*, 40 were *P. vivax*, and 11 showed mixed infections with both species. To evaluate the extract-free workflow, 5 µL of whole blood from each specimen was subjected to Triton X-100-based ambient lysis, and the resulting crude lysate was directly introduced into the one-pot RPA-CRISPR reaction. In parallel, 100 µL aliquots from each sample underwent conventional DNA extraction to enable comparative evaluation by reference real-time PCR and by the RPA-CRISPR assay using purified DNA input.

### Reference PCR Testing and Extracted-DNA Comparison

Genomic DNA was extracted from whole blood samples using the EZ1 DSP DNA Blood Kit on the EZ1 Advanced XL instrument (Qiagen GmbH, Hilden, Germany) as previously described [9-14]. DNA was eluted in 50 µL of hydration buffer and either used immediately or stored at −20 °C until analysis. Reference testing was performed using a validated quantitative PCR (qPCR) assay targeting the conserved *Plasmodium* 18S rRNA gene, with species-specific probes for discrimination of *P. falciparum* and *P. vivax* ^27^. The PCR results served as the clinical reference standard for determination of diagnostic sensitivity, specificity, and overall concordance. Parasite burden in PCR-positive samples was estimated based on the cycle threshold (Ct) values. Samples were stratified into three categories: high parasite burden (Ct <25), moderate parasite burden (Ct 25-30), and low parasite burden (Ct >30). The diagnostic performance of the extract-free one-pot RPA-CRISPR assay was subsequently evaluated within each parasite burden category.

Furthermore, purified DNA obtained through the extraction protocol was also evaluated using the RPA-CRISPR assay under identical reaction conditions to those applied in the extract-free workflow. This parallel testing enabled direct comparison of crude lysate and column-purified DNA within the same RPA-CRISPR platform, allowing evaluation of matrix effects, potential enzyme inhibition, and performance trade-offs associated with eliminating DNA purification.

### Statistical Analysis

A total of 116 PCR-positive cases and 109 PCR-negative controls were included to ensure adequate precision of diagnostic performance estimates. Assuming an expected sensitivity and specificity of 95%, this sample size provides approximately ±5% precision for two-sided 95% confidence intervals and >90% power to detect a ≥10% difference in sensitivity compared with reference PCR (α = 0.05). The analytical limit of detection (LOD) was determined using serial dilutions of quantified control DNA and defined as the lowest concentration detected in ≥95% of replicates. Detection probabilities were modeled using probit regression analysis. Diagnostic accuracy metrics, including sensitivity and specificity, were calculated with exact (Clopper-Pearson) 95% confidence intervals. Agreement between the one-pot RPA-CRISPR assay and the reference PCR method was assessed using Cohen’s kappa coefficient. To evaluate the relationship between parasite burden and assay performance, logistic regression analysis was performed to assess the association between PCR Ct values and the probability of detection by the RPA-CRISPR assay. Differences in detection rates across parasite burden categories were evaluated using χ^2^ or Fisher’s exact tests, as appropriate. Continuous variables were analyzed using two-sided t-tests or appropriate nonparametric equivalents, with p < 0.05 considered statistically significant. All statistical analyses were performed using Python (v3.11; SciPy and statsmodels) and R (v4.3.2).

## RESULTS

### Analytical Specificity of the One-Pot Ambient RPA-CRISPR

We first evaluated the analytical specificity of the ambient one-pot RPA-CRISPR system for discriminating between *P. falciparum* and *P. vivax* using quantified control DNA templates at 1 × 10^3^ copies/µL. Species-specific RPA primers and corresponding crRNAs targeting conserved regions within the 18S rRNA gene were assessed under fully ambient conditions (25 °C) in the single-tube format. Robust and distinct test-band formation was observed exclusively in reactions containing the matching target DNA, whereas no detectable signal was observed when non-matching species templates were tested under identical conditions (Figure 2). Negative template controls consistently showed absence of test-band development, confirming lack of nonspecific amplification or reporter cleavage. Importantly, no cross-reactivity was detected between *P. falciparum* and *P. vivax* targets, demonstrating high analytical specificity of both the RPA primer sets and CRISPR-crRNA complexes. These findings indicate that species-specific detection is preserved under room-temperature, single-tube reaction conditions and support the feasibility of selective molecular identification using the ambient RPA-CRISPR platform.

**Figure 1.**
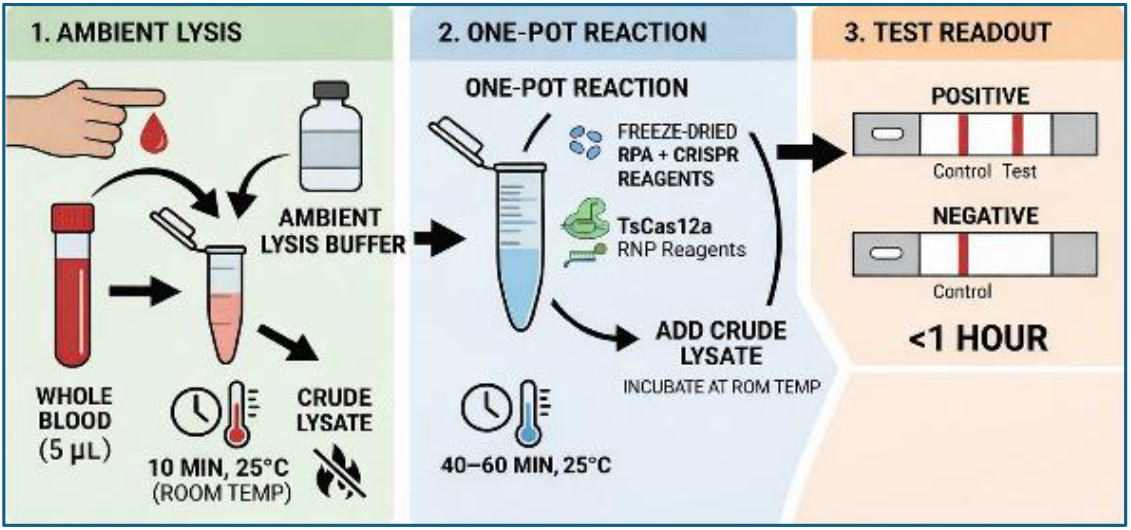
Workflow of the one-pot, ambient-temperature malaria diagnostic platform. Raw blood is lysed at room temperature using an extract-free lysis buffer to generate crude lysate. The lysate is directly added to a single reaction tube containing RPA and CRISPR reagents for simultaneous amplification and detection at room temperature. Results are visualized by lateral flow assay, with the entire process completed under one hour without electricity or specialized equipment.

**Figure 2.**
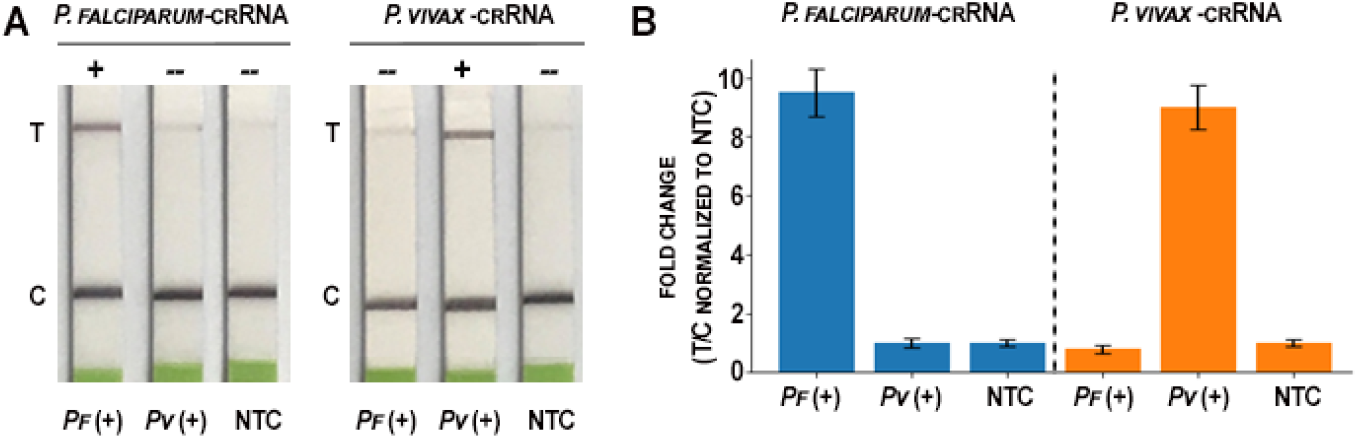
Species-specific detection of *Plasmodium* using the one-pot ambient RPA-CRISPR assay with lateral flow readout. (A) Representative lateral-flow strips demonstrating species-specific detection using CRISPR-Cas12a with guide RNAs targeting *P. falciparum* or *P. vivax*. Using the *P. falciparum*-specific crRNA (left), a visible test line (T) is observed only in *P. falciparum*-positive samples (Pf+), with no signal detected in *P. vivax*-positive samples (Pv+) or no-template controls (NTC). Using the *P. vivax*-specific crRNA (right), the test line appears exclusively in *P. vivax*-positive samples, with no cross-reactivity observed in *P. falciparum* samples or NTC. The control line (C) confirms proper strip function. (B) Quantitative densitometric analysis of the lateral-flow test lines. Test-line intensities were normalized to the corresponding control-line intensities (T/C ratio) and expressed as fold change relative to the negative control (NTC). Bars represent mean ± SD.

### Analytical Sensitivity of the One-Pot Ambient RPA-CRISPR

Analytical sensitivity of the one-pot RPA-CRISPR assay was evaluated at 25 °C using ten-fold serial dilutions of ddPCR-quantified *P. falciparum* and *P. vivax* genomic DNA prepared in nuclease-free buffer, ranging from 1 × 10^4^ to 1 × 10^0^ copies/µL. A conventional two-step RPA-CRISPR detection workflow was performed in parallel using the identical dilution series to enable controlled head-to-head comparison of detection kinetics and sensitivity thresholds. The LOD of the one-pot ambient RPA-CRISPR assay, defined as the lowest concentration detected in ≥95% of replicate reactions, was 10 copies/µL for both *P. falciparum* and *P. vivax* (Figure 3). Probit regression analysis confirmed a ≥95% detection probability at the established LOD for both targets (Figure 3). Therefore, the one-pot ambient RPA-CRISPR assay reliably detected *P. falciparum* and *P. vivax* at concentrations as low as 10 copies/µL, demonstrating strong analytical sensitivity. No false-positive test bands were observed among negative template controls (NTCs). Extension of incubation time beyond 60 minutes did not significantly improve the LOD. The conventional two-step RPA-CRISPR assay demonstrated comparable analytical performance to the one-pot format (Figure 4), confirming that integration into a single-tube workflow does not compromise sensitivity. These findings indicate that the one-pot RPA-CRISPR assay maintains high analytical sensitivity under ambient conditions, supporting its suitability for room-temperature molecular detection without active temperature regulation.

**Figure 3.**
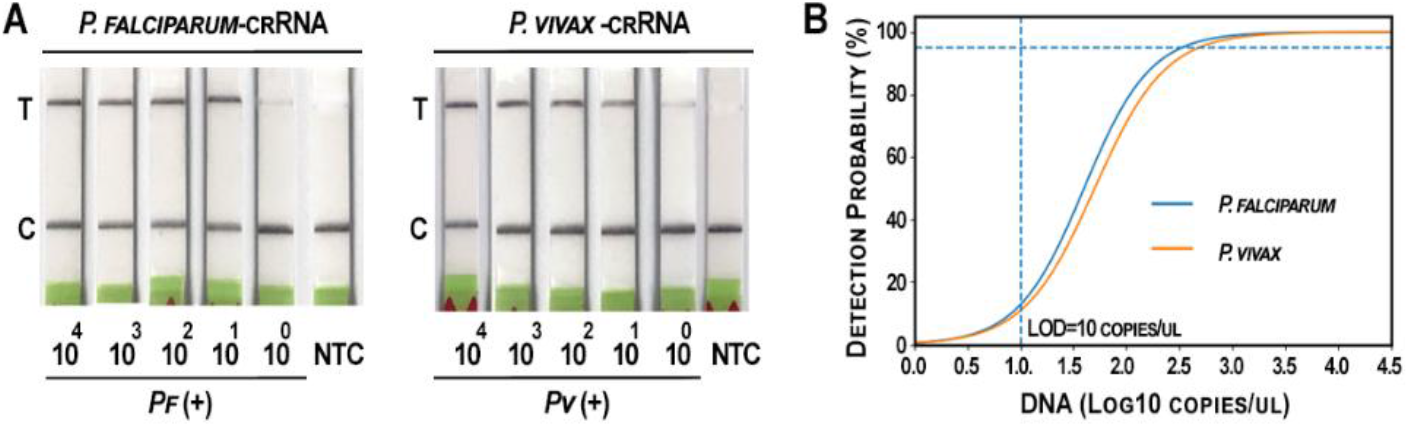
Analytical sensitivity of the one-pot ambient RPA-CRISPR assay. (A) Lateral-flow detection of ten-fold serial dilutions of *P. falciparum* and *P. vivax* genomic DNA (10^4^–10^0^ copies/µL) at 25 °C using species-specific crRNAs. NTC, no-template control. The limit of detection (LOD) was 10 copies/µL for both species. (B) Detection probability as a function of log_10_ DNA concentration determined by probit regression analysis of replicate experiments. The dashed horizontal line indicates the 95% detection probability, and the dashed vertical line marks the LOD (10 copies/µL).

**Figure 4.**
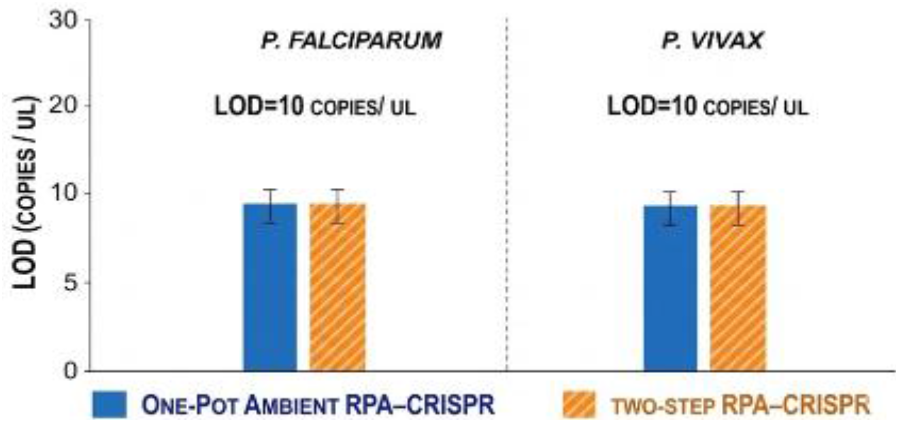
Comparison of Analytical Sensitivity Between One-Pot and Two-Step RPA-CRISPR Assays. LOD comparison for *P. falciparum* and *P. vivax* using the one-pot ambient RPA-CRISPR assay (blue bars) and the conventional two-step RPA-CRISPR assay (orange, hatched bars). The LOD was 10 copies/µL for both species in both assay formats. Data represents the mean of three independent experiments; error bars indicate SD. No significant difference in analytical sensitivity was observed between the two methods for either species (All p>0.05).

### Analytical Performance of Extract-Free One-Pot Ambient RPA-CRISPR

Serial dilutions of *P. falciparum* and *P. vivax* genomic DNA were spiked into malaria-negative whole blood. The spiked whole-blood samples were divided into two equal portions for a head-to-head comparison of processing methods. The first portion was subjected directly into the extract-free one-pot RPA-CRISPR reaction. The second portion was subjected to standardized column-based DNA isolation, and the purified DNA was analyzed using both the one-pot RPA-CRISPR assay and a reference multiplex real-time PCR. Using purified DNA, both the RPA-CRISPR assay and the reference real-time PCR assay demonstrated an identical limit of detection of 10 copies/µL for *P. falciparum* and *P. vivax*, consistent with previous report ^28^. The extract-free RPA-CRISPR format, utilizing the crude lysates, achieved an LOD of 100 copies/µL for both species (Figure 6). Furthermore, no cross-reactivity was observed in any assay format when non-target templates were tested, confirming 100% analytical specificity across all platforms. Compared with RPA-CRISPR and PCR using purified DNA samples (LOD: 10 copies/µL), the extract-free one-pot ambient RPA-CRISPR assay showed lower sensitivity with an LOD of 100 copies/µL.

**Figure 5.**
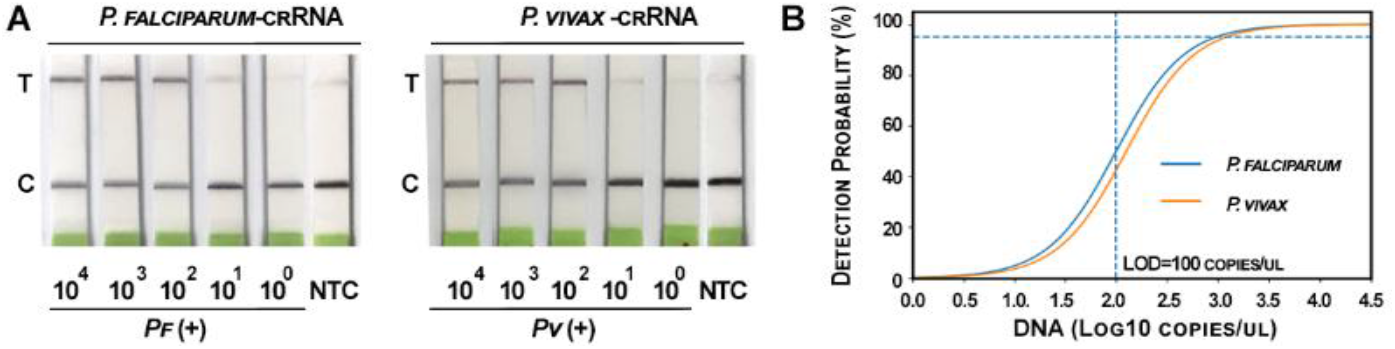
Analytical sensitivity of the extract-free one-pot ambient RPA-CRISPR assay. (A) Lateral-flow detection of ten-fold serial dilutions of *P. falciparum* and *P. vivax* genomic DNA (10^4^-10^0^ copies/µL) spiked into malaria-negative blood and processed by Triton X-100-based ambient lysis prior to one-pot RPA-CRISPR detection at 25 °C. NTC, no-template control. The LOD was 100 copies/µL for both species. (B) Probit regression analysis showing detection probability versus log_10_ DNA concentration. The dashed horizontal line indicates 95% detection probability, and the dashed vertical line marks the LOD (100 copies/µL).

**Figure 6.**
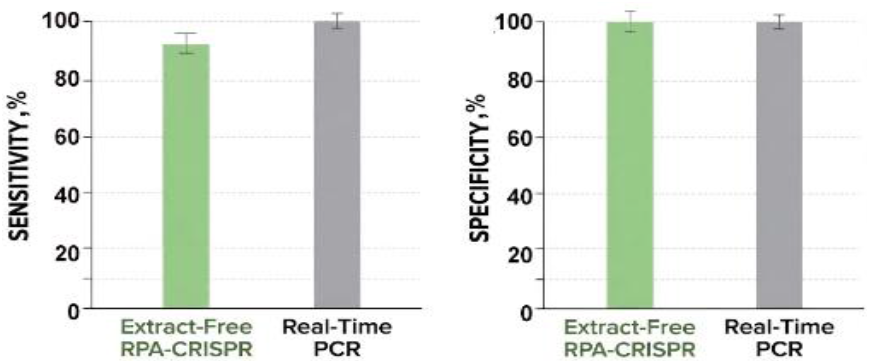
Diagnostic performance of the extract-free one-pot RPA-CRISPR assay compared with real-time PCR in the clinical specimens. Sensitivity and specificity of the extract-free one-pot RPA-CRISPR assay were evaluated using realtime PCR as the reference standard. Of the 116 PCR-positive samples, 65 were identified as *Plasmodium falciparum*, 40 as *P. vivax*, and 11 as mixed infections. The extract-free one-pot RPA-CRISPR assay detected 61/65 *P. falciparum*, 37/40 *P. vivax*, and 10/11 mixed infections, while all 109 PCR-negative samples were correctly identified as negative. Overall sensitivity and specificity were 93.1% (108/116) and 100% (109/109), respectively. Error bars represent 95% confidence intervals (Clopper-Pearson method).

### Diagnostic Performance of the Extract-Free Ambient One-Pot RPA-CRISPR Assay in Clinical Specimens

The extract-free ambient one-pot RPA-CRISPR assay was evaluated in a clinical cohort comprising 116 malaria-positive cases and 109 negative controls. Parasite burden among PCR-positive cases was estimated using Ct values of qPCR. Of the 116 positive cases, 46 (39.7%) exhibited high parasite density (Ct <25), 41 (35.3%) moderate density (Ct 25-30), and 29 (25.0%) low density (Ct >30) (Supplementary Table 2). High parasite burden was more frequently observed in *P. falciparum* infections, whereas *P. vivax* cases were more evenly distributed between moderate and low parasite density categories. Among the 116 PCR-positive samples, the extract-free one-pot RPA-CRISPR assay detected 61 of 65 *P. falciparum* cases, 37 of 40 *P. vivax* cases, and 10 of 11 mixed infections, while all 109 PCR-negative samples tested negative. Overall, the assay achieved a sensitivity of 93.1% (108/116) and specificity of 100% (109/109) (Figure 8). Stratified analysis showed that sensitivity of the extract-free RPA-CRISPR assay correlated with parasite burden (Supplementary Table 3). Among samples with high parasite density (Ct <25), the assay detected 45 of 46 cases (97.8%). Sensitivity was 95.1% (39/41) for moderate parasite density (Ct 25-30) and 82.8% (24/29) for low-density samples (Ct >30). Logistic regression analysis further confirmed a significant association between parasite burden (PCR Ct value) and the probability of detection by the RPA-CRISPR assay, with lower Ct values (higher parasite density) associated with increased likelihood of detection (*p* < 0.05). Furthermore, agreement between the extract-free one-pot RPA-CRISPR assay and the reference qPCR assay was evaluated using Cohen’s κ statistic and McNemar’s test. Overall agreement between the two methods was 96.4% (217/225), with a κ value of 0.93 indicating near-perfect agreement. McNemar’s test identified a small but statistically significant difference between the assays (*p* = 0.008) (Supplementary Table 4), reflecting eight samples that were positive by qPCR but negative by the RPA-CRISPR assay. Sensitivity: 93.1% (108/116), Specificity: 100% (109/109), Overall agreement: 96.4% (217/225), Cohen’s κ (kappa): 0.93, McNemar’s test: *p* = 0.008. Agreement between the RPA-CRISPR assay and qPCR was 96.4% (κ = 0.93). McNemar’s test indicated a small but significant difference between the methods (p = 0.008), reflecting eight qPCR-positive samples not detected by RPA-CRISPR. The modest reduction in diagnostic sensitivity compared with PCR is consistent with the approximately one-log decrease in analytical sensitivity observed in the matrix-spiking experiments and likely reflects inhibitory effects associated with crude whole-blood matrices.

## Discussion

In this study, we developed an extract-free, ambient-temperature, one-pot RPA-CRISPR diagnostic platform capable of detecting *Plasmodium* species directly from whole blood. By integrating detergent-based lysis with simultaneous isothermal amplification and CRISPR detection in a single closed-tube reaction, the assay eliminates nucleic acid purification, thermal cycling, and specialized instrumentation. This simplified workflow enables molecular detection under ambient conditions and may facilitate deployment in decentralized or resource-limited settings.

Analytical evaluation demonstrated a LOD of 100 copies/µL using crude whole-blood lysates, whereas the RPA-CRISPR chemistry achieved 10 copies/µL when purified DNA was used. The approximately one-log reduction in sensitivity likely reflects inhibitory components present in whole blood rather than intrinsic limitations of the amplification-detection chemistry. Despite this reduction, the assay maintained high analytical specificity and showed strong concordance with reference PCR. Clinical evaluation using real-time PCR as the reference standard showed strong concordance, with the extract-free workflow achieving 93.1% sensitivity and 100% specificity. Furthermore, diagnostic sensitivity varied with parasite burden, remaining high in samples with moderate to high parasite density but decreasing in low-density infections.

Because the *Plasmodium* 18S rRNA gene is present in multiple copies per parasite genome, the analytical LOD of 100 copies/µL corresponds approximately to 12-20 parasites/µL, a range relevant for molecular malaria diagnostics (Supplementary Table 5). Compared with PCR-based methods, the extract-free one-pot RPA-CRISPR platform offers several operational advantages. These particularly include room-temperature processing, elimination of nucleic acid extraction, rapid turnaround, minimal instrumentation, and low reagent cost (∼$15 per test). Together, these features support its potential use for POC malaria diagnosis in resource-limited settings (Supplementary Table 6).

Several limitations should be considered. This study represents a pilot feasibility evaluation with a modest sample size; larger multicenter studies are needed to more precisely define diagnostic performance across diverse epidemiological settings. The analysis focused on *P. falciparum* and *P. vivax*, and future studies should evaluate additional *Plasmodium* species and more complex mixed infections to broaden clinical applicability. The sensitivity of the extract-free one-pot RPA-CRISPR assay in crude whole blood was slightly lower than that of real-time PCR, suggesting that further optimization of reaction chemistry may improve detection in low-parasitemia samples. Although the assay is designed for POC deployment and operates under ambient conditions without nucleic acid extraction or specialized instrumentation, the current evaluation was performed under controlled laboratory conditions using archived specimens. Field-based validation in endemic settings will therefore be essential to assess performance under real-world conditions, including variations in temperature, sample handling, and operator training. In addition, systematic studies of reagent stability, interference, and dilution-recovery in crude biological matrices will be needed to further characterize assay robustness and support deployment in resource-limited environments.

In summary, the ambient one-pot RPA-CRISPR platform combines molecular-level sensitivity with a simplified workflow that eliminates nucleic acid extraction and temperature-controlled instrumentation. The assay demonstrates high diagnostic accuracy in clinical samples and operates entirely under ambient conditions with a rapid turnaround time. These features suggest that the platform may provide a practical molecular diagnostic approach for malaria detection in decentralized or resource-limited settings.

## Data Availability

All data produced in the present study are available upon reasonable request to the authors

## Ethics Approval and Consent to Participate

The study protocol (IRB 231876) was reviewed and approved by Jiangsu Province Hospital of Chinese Medicine. All procedures were conducted under institutional review board-approved protocol, following the ethical standards. Written informed consent was obtained from all participants prior to sample collection and study enrollment.

## CRediT authorship contribution statement

Jipei Liao: Conceptualization, Methodology, Investigation, Data curation, Formal analysis, Visualization, Writing – original draft. Yun Su: Investigation, Resources, Sample collection, Validation, Writing – review & editing. Feng Jiang: Conceptualization, Supervision, Project administration, Funding acquisition, Writing – review & editing.

## Acknowledgements

The authors wish to thank the Biostatistics Shared Service at the University of Maryland Marlene and Stewart Greenebaum Cancer Center for their invaluable contribution in conducting the statistical analysis for this study.

